# Humoral and cellular immune responses upon SARS-CoV-2 vaccines in patients with anti-CD20 therapies: A systematic review and meta-analysis of 1342 patients

**DOI:** 10.1101/2021.09.30.21264335

**Authors:** Simeon Schietzel, Manuel A. Anderegg, Andreas Limacher, Alexander Born, Michael P. Horn, Britta Maurer, Cédric Hirzel, Daniel Sidler, Matthias B. Moor

## Abstract

**Background:** Immune responses upon SARS-CoV-2 vaccination in patients receiving anti-CD20 therapies are impaired but vary considerably. We conducted a systematic review and meta-analysis of the literature on SARS-CoV-2 vaccine induced humoral and cell-mediated immune response in patients previously treated with anti-CD20 antibodies.

**Methods:** We searched PubMed, EMBASE, Medrxiv and SSRN using variations of search terms “anti-CD20”, “vaccine” and “COVID” and included original studies up to August 21^st^,2021. We excluded studies with missing data on humoral or cell-mediated immune response, unspecified methodology of response testing, unspecified timeframes between vaccination and blood sampling or low number of participants (≤ 3). We excluded individual patients with prior SARS-CoV-2 infection or incomplete vaccine courses. Primary endpoints were humoral and cell-mediated immune response rates. Pre-specified subgroups were time of vaccination after anti-CD20 therapy (< vs > 6 months), time point of response testing after vaccination (< vs > 4 weeks) and disease entity (autoimmune vs cancer vs renal transplant). We used random-effects models of proportions.

**Findings:** Ninety studies were assessed. Inclusion criteria were met by 23 studies comprising 1342 patients. Overall rate of humoral response was 0.41 (95% CI 0.35 – 0.47). Overall rate of cell-mediated immune responses was 0.71 (95% CI 0.47 – 0.90). Longer time interval since last anti-CD20 therapy was associated with higher humoral response rates > 6 months 0.63 (95% CI 0.53 – 0.72) vs < 6 months 0.2 (95% CI 0.03 – 0.43); p = 0.001. Compared to patients with haematological malignancies or autoimmune diseases, anti-CD20 treated kidney transplant recipients showed the lowest vaccination response rates.

**Interpretation:** Patients on anti-CD20 therapies can develop humoral and cell-mediated immune responses after SARS-CoV-2 vaccination, but subgroups such as kidney transplant recipients or those with very recent B-cell depleting therapy are at high risk for non-seroconversion and should be individually assessed for personalized SARS-CoV-2 vaccination strategies. Potential limitations are small patient numbers, heterogeneous diseases and assays used.

**Funding:** This study was funded by Bern University Hospital.

## INTRODUCTION

The severe impact of the coronavirus disease 19 (COVID-19) pandemic has led to the implementation of worldwide vaccination programs.. Even though SARS-CoV2 vaccines have been made widely available, immunocompromised patients may still be at significant risk for severe COVID-19 after immunization. B-cell depleting therapy in particular is associated with impaired vaccination responses, as already demonstrated in pre-pandemic studies ^1–3^. In addition, disease entities and patient factors, such as individual and disease-specific B-cell repopulation kinetics further influence response rates (4,5). Also, an adequate time interval between anti-CD20 therapy and vaccination seems crucial as previously demonstrated by immune response rates upon influenza vaccines ^4^.

With the broad availability of SARS-CoV2 vaccines in many countries, strategies aimed at understanding and improving the immunogenicity of vaccines are urgently needed for patients undergoing anti-CD20 therapy. We therefore performed a systematic review and meta-analysis of humoral and cell-mediated immune responses after administration of SARS-CoV2 vaccines in patients treated with anti-CD20 antibodies focusing on quantitative measures, diseases entities and duration since last anti-CD20 therapy.

## METHODS

We performed a systematic review and meta-analysis of peer-reviewed studies and pre-prints available online and reported it according to the Preferred Reporting Items for Systematic Reviews and Meta-Analyses ^5^.

### Definitions

We defined anti-CD20 therapy as treatment with rituximab, rituximab-abbs, rituximab-arrx, rituximab-hyaluronidase, rituximab-pvvr, ocrelizumab, obinutuzumab, ofatumumab and ibritumomab tiuxetan. We defined rituximab as monotherapy if explicitly reported. In most of the included studies however, it was not defined if anti-CD20 treatment was administered as monotherapy or in combination with other immunosuppressives. In these cases, we assumed concomitant immunosuppressive co-medication as disease types and enumerated baseline medication highly suggested anti-CD20 therapy being part of an immunosuppressive combination therapy.

We defined SARS-CoV-2 vaccine elicited humoral immune response as detection of anti-spike antibodies (anti-RBD or anti-S1 (spike protein) SARS-CoV2) above the cut-off reported by the manufacturer of the given assay.Vaccine elicited cell-mediated immune response was defined as detection of SARS-CoV-2 specific T-cells either measured by, T-EliSpot ^6–9^, interferon-γ release assays ^10,11^ or activation-induced marker (AIM) detection ^12,13^ in flow cytometry-sorted cells. AIM used for the detection of vaccine elicited T-cells response were CD4+CXCR5+PD1+ and CD38+HLA-DR+ ^12^ as well as S-specific OX40+ 41-BB+ CD4+ and CD69+ 41BB+ CD8+ ^13^.

Autoimmune diseases were defined as a collective of diseases characterized by aberrant immune responses including the presence of anti-bodies or T cells reacting with self-antigens that are treated with immunosuppressants.

### Eligibility criteria

We considered all original research studies that investigated serologic and/or cell mediated responses upon SARS-CoV-2 vaccination in patients with anti-CD20 therapies potentially eligible for inclusion. Pre-specified exclusion criteria were exclusive focus on participants with previous COVID-19 infection or incomplete vaccination schedules, unspecified time frames between vaccination and blood sampling, unspecified methodology for detection of antibody- or cell-mediated immunity (specification of manufactures and detection kits mandatory), number of investigated participants lower or equal than 3, missing numbers of positive versus negative humoral or cell-mediated immune responders. In addition, review and guideline articles as well as all search results not meeting the topic of our research question were excluded.

### Information sources and search strategy

PubMed (up to August 21, 2021), EMBASE (up to August 21, 2021), as well as pre-print servers medrxiv, SSRN and SSRN-Lancet (January 01, 2020 to August 21, 2021) were accessed online and searched within title/abstract without language restrictions.

For PubMed ^14^, a search for “rituximab OR anti-cd20 AND covid AND vaccine” within title/abstract was performed. For EMBASE ^15^, a search for “rituximab OR anti-CD20 AND covid AND vaccine” within title/abstract was performed. For medrxiv ^16^, a search for “rituximab AND covid AND vaccine” was performed. Additionally, a search for “anti-CD20 AND covid AND vaccine” was performed. A third search using “anti-CD20 AND covid19 AND vaccine” was performed. For SSRN ^17^, a search for “rituximab AND covid AND vaccine” was performed. Another search using terms “anti-cd20 AND covid AND vaccine” was performed. For SSRN Preprints with The Lancet ^18^, a search for “rituximab AND covid AND vaccine” was performed. Another search for “anti-CD20 AND covid AND vaccine” was performed.

### Selection process

We executed the process of studies selection in accordance with Cochrane recommendations ^19^. Two authors (SS and MA) independently assessed all search results of PubMed database, and two authors (SS and MBM) independently assessed all search results of EMBASE database and preprint servers. In cases of divergent selections, a third author (MA or MBM, respectively) was consulted. Fulfilments of in- and exclusion criteria were reviewed again by each of the three authors and decision processes were mutually rechecked and discussed thereafter. Discrepancies could be unequivocally resolved in all cases with full agreement by all authors. We did not need to apply the pre-specified mode of majority decisions due to persistent disagreements. We did not apply automation tools.

### Data collection process

Tabular and text data of study population subsets with a history of anti-CD20 therapies were manually copied and independently downloaded by each reviewer. Extraction of graphical figure data was performed by image analysis in selected cases. We did not apply automation tools.

### Data items

Next to the inclusion and exclusion criteria specified above, we extracted the following information from the searched studies:

#### Primary outcome data

Percentage of participants (with anti-CD20 therapies, no history of previous COVID19 disease, complete SARS-CoV-2 vaccination course) with positive serologic and/or cell-mediated immune response after SARS-CoV-2-vaccination.

#### Data for subgroup generation

Disease types of study population, type of immunosuppressive therapy (anti-CD20 +/- other immunosuppressive treatment), primary outcome data separated by time since last anti-CD20 treatment (before vs after 6 months) as well as by timing of response testing (<4 weeks vs >4 weeks after final SARS-CoV-2 vaccination).

#### Data for quality evaluation

Study design, method of cell-mediated immune (CMI) response measurement, manufactures of detection kits and respective cut-off values for test positivity, title of study, digital object identifier (DOI) and PubMed identifier (PMID) for repeated duplication checks. In cases where cut-off values of a manufacturer’s kit for antibody or CMI response testing were not specified in the methods section, we retrieved these data from the manufacturer’s website.

### Risk of bias assessment

We manually assessed the risk of bias of included studies using the Newcastle-Ottawa Scale (NOS) for assessing the quality of non-randomised studies in meta-analyses by Wells et al. ^20^. Three investigators (SS, AB and MBM) independently assigned a number of quality criteria (minimum 0, maximum 9) for each study. A fourth investigator (MA) summarised results according to a pre-specified mode of majority decision. Threshold of an optimal follow-up period was estimated as at least 4 weeks after completed vaccination ^21^.

### Effect measures

For both outcomes of humoral and cell-mediated immunity, number and proportion of responders was used in the synthesis and presentation of results.

### Synthesis methods

Synthesis was first obtained by tabulating the studies using Microsoft Excel and comparing against a list of exclusion criteria. No missing data were present in the included studies. We performed a random-effects meta-analysis of proportions using the method of Der Simonian & Laird, with pooled estimates calculated by Freeman-Tukey Double Arcsine Transformation^22^ to stabilize the variances. Statistical heterogeneity was quantified using the I-squared measure, taken from the inverse-variance fixed-effect model ^23^. For individual studies, the Wilson score 95% confidence interval is displayed. To explore possible causes of heterogeneity between studies, we performed pre-specified subgroup analyses specified above.

Quantitative analyses and graphical displays were done in Stata version 17 using the command metaprop version 10.1. Meta-regression and small study effects analysis was conducted in R version 4.0.5 (meta-package 4.19-1). No sensitivity analyses were performed.

### Reporting bias assessment

Small study effects were assessed by a funnel plot and a regression test for funnel-plot asymmetry ^24^.

### Certainty assessment

No procedures were performed to assess confidence in the body of evidence.

## RESULTS

### Search results

The study selection process is presented in detail in **Figure 1**. Searches in PubMed ^14^ and EMBASE (12) yielded 73 and 39 results respectively. Searches within pre-print servers yielded 12 studies from medrxiv ^16^ and 23 studies from SSRN ^17^ and SSRN Preprints with The Lancet ^18^. After removal of 57 duplicates, 90 studies remained for eligibility assessment. Title and abstract screening led to exclusion of 56 additional articles. Full-text screening of the remaining 34 articles then led to further exclusion of 11 articles **(Supplementary Appendix, pages 3-8)**. A total of 23 studies with data of 1342 participants fully met inclusion criteria and were eventually included in the Meta-Analysis.

**Figure 1.**
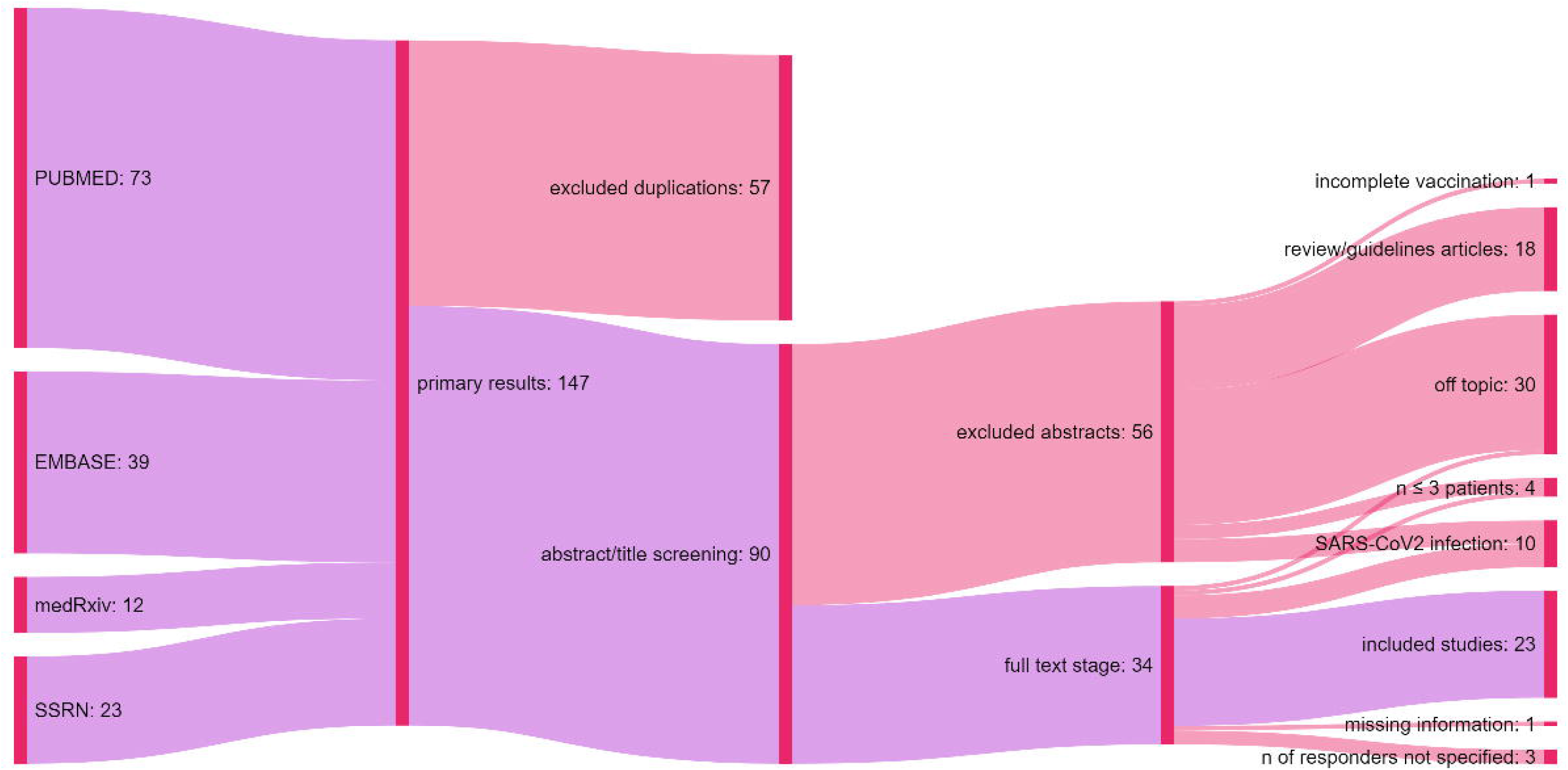
Flowchart describing the study search and selection process.

### Study characteristics

Study characteristics of the included publications ^6–13,25–39^ are summarized in **Table 1**. Information to assay details of antibody detection and CMI are indicated in **Supplementary Appendix, pp. 8-9**.

**Table 1:**
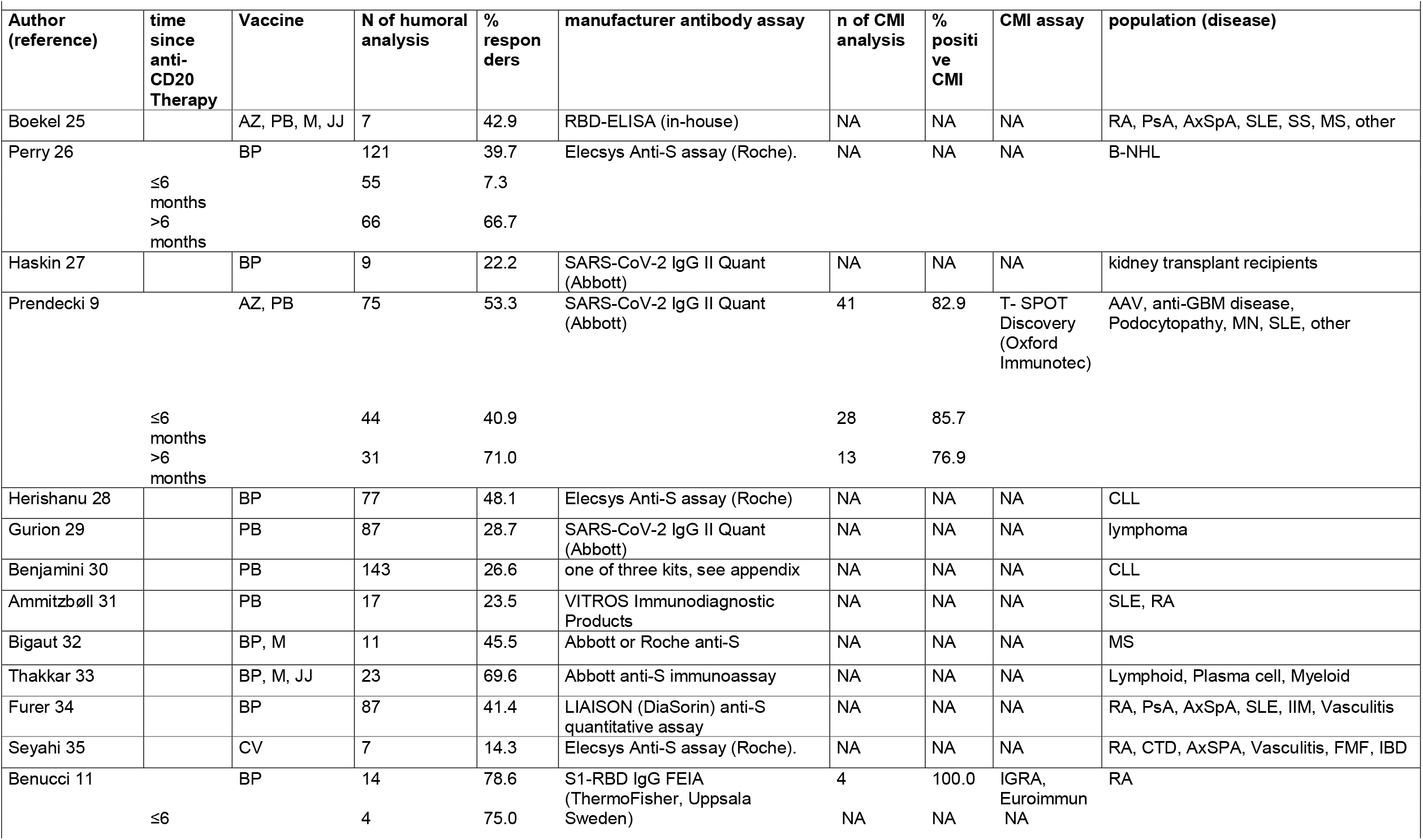
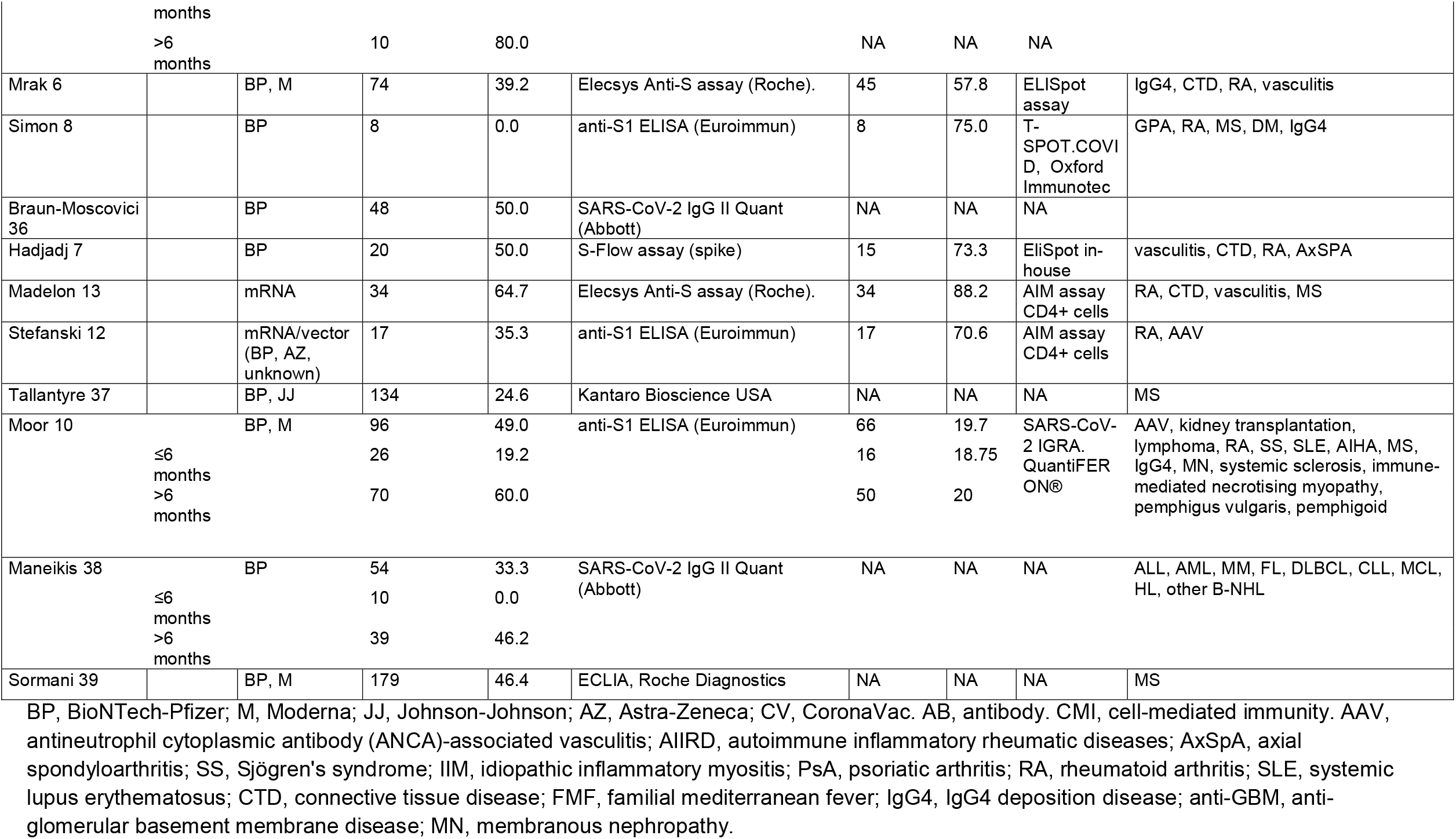
List of included studies.

### Risk of bias

**Supplemental Appendix, p. 10** shows the results of the risk of bias assessment for the individual studies included. Applying the Newcastle-Ottawa Scale (NOS) 8 out of 9 possible quality rating criteria could be met (“Adequacy of follow-up” not applicable). Risk of bias ratings for the 23 included studies were low to moderate.

### Results of individual studies

Details regarding the calculations of proportions for meta-analysis using a random-effects model calculation of the effect size (ES) are mentioned in the methods section. Pooled estimates of proportions are shown, with 95% confidence interval. The last column shows the weight of the specific study in percentage.

### Humoral response to vaccination

Humoral responses were highly heterogeneous with a rate of responders ranging from 0 to about 80%, resulting in an overall humoral response rate of 41%. An I^2 of 74% confirmed heterogeneity in this study collection **(Figure 2)**. Therefore, studies were subjected to pre-specified sub-group analyses including time since last anti-CD20 therapy, indication, and time of post-vaccination testing.

**Figure 2.**
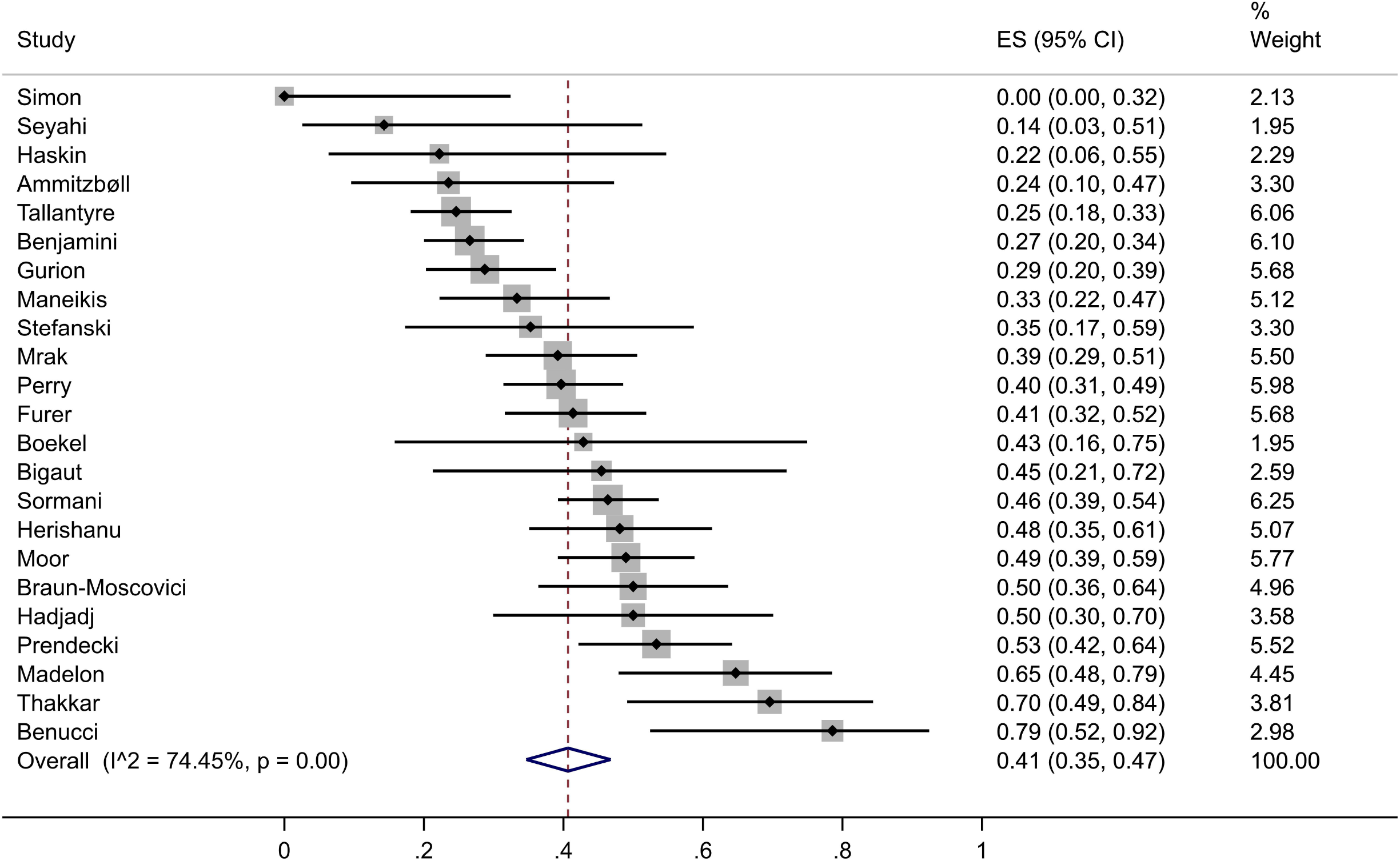
Humoral immune responses across all included studies. ES, effect size. CI, confidence interval.

Stratification based on a 6 months threshold since last anti-CD20 therapy showed that studies with shorter intervals reported significantly lower numbers of responders (20% vs. 63%) with exception for one study including exclusively patients with rheumatoid arthritis **(Figure 3)**. Benucci et al. reported a markedly higher humoral response rate of 75% compared with other short-term studies, which probably accounts for the high heterogeneity of this subgroup.

**Figure 3.**
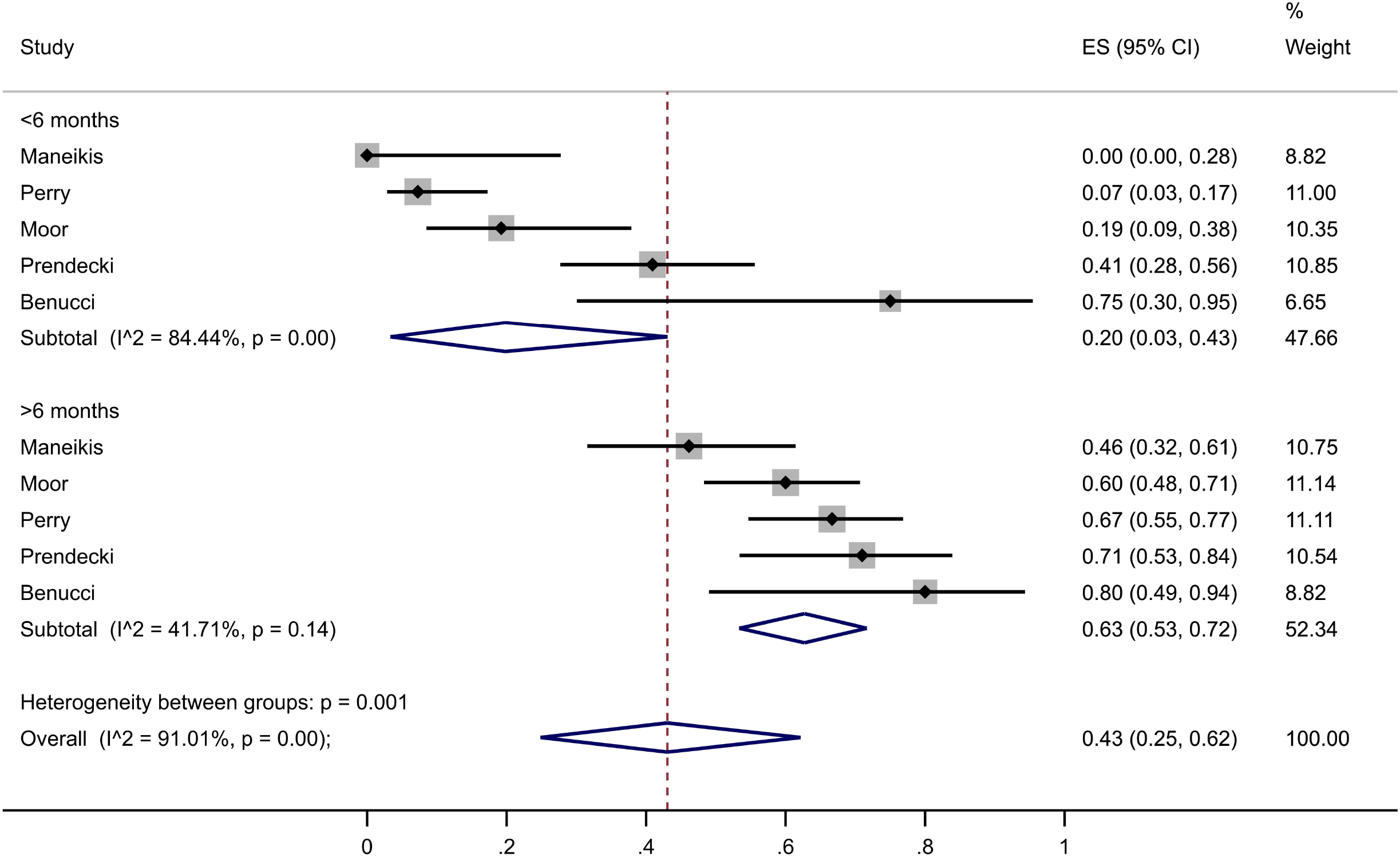
Humoral immune responses according to pre-specified subgroups of <6 or >6 months of time since the last dose of anti-CD20 therapy. ES, effect size. CI, confidence interval.

Analysis of humoral response by indication for B-cell-depletion (autoimmune diseases, haematological malignancy, and kidney transplantation) is shown in **Figure 4**. Pooled humoral response rates were similar for autoimmune diseases and cancer (43% vs 37%), but markedly lower for patients having undergone kidney transplantation (14%).

**Figure 4.**
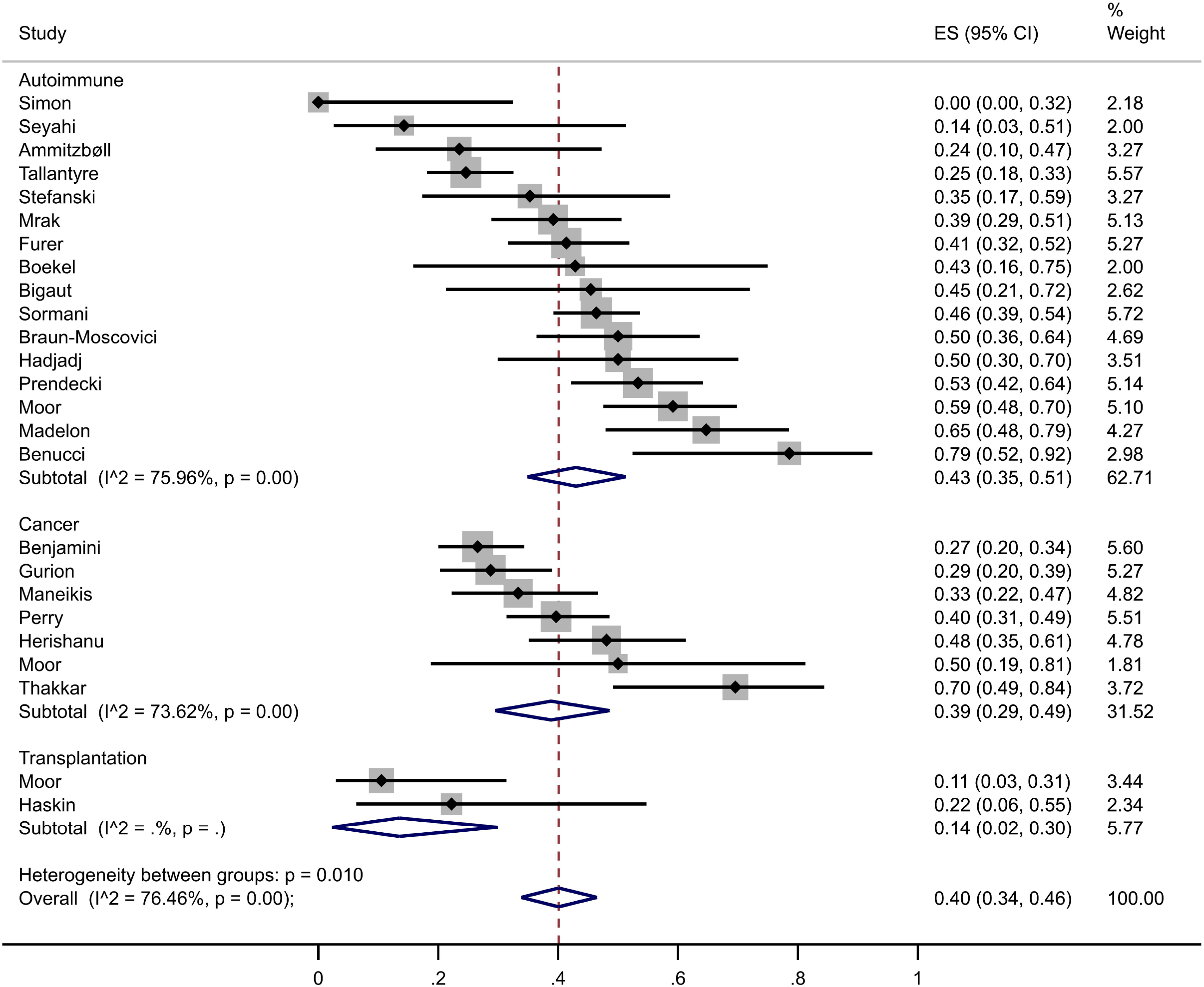
Humoral immune responses according to pre-specified subgroups of indications for anti-CD20 therapy. Cancer indicates hematological malignancy. CI, confidence interval.

**Supplementary appendix p.2** shows humoral response stratified by time since vaccination based on a 4-week threshold. Humoral response (pooled response rate) was 45% for > 4 weeks post vaccination versus 36% for 0-4 weeks post vaccination. In a meta-regression for humoral immune responses according to population-level mean or median of time between last vaccine and antibody measurement, the antibody response tended to increase by 3.9% per 10-day increase in time since vaccination. The effect was not significant (risk difference: 3.9%, 95%CI −1.2% to 9.1%, p= 0.136).

### Cell-mediated immune response to vaccination

Cell-mediated immune response rates (not stratified, including EliSpot, IGRA and AIM) varied from 20% to 100%, with an overall pooled response rate of 71% **(Figure 5)**. The heterogeneity of the included studies is large as indicated by an I^2 of 90.85%.

**Figure 5.**
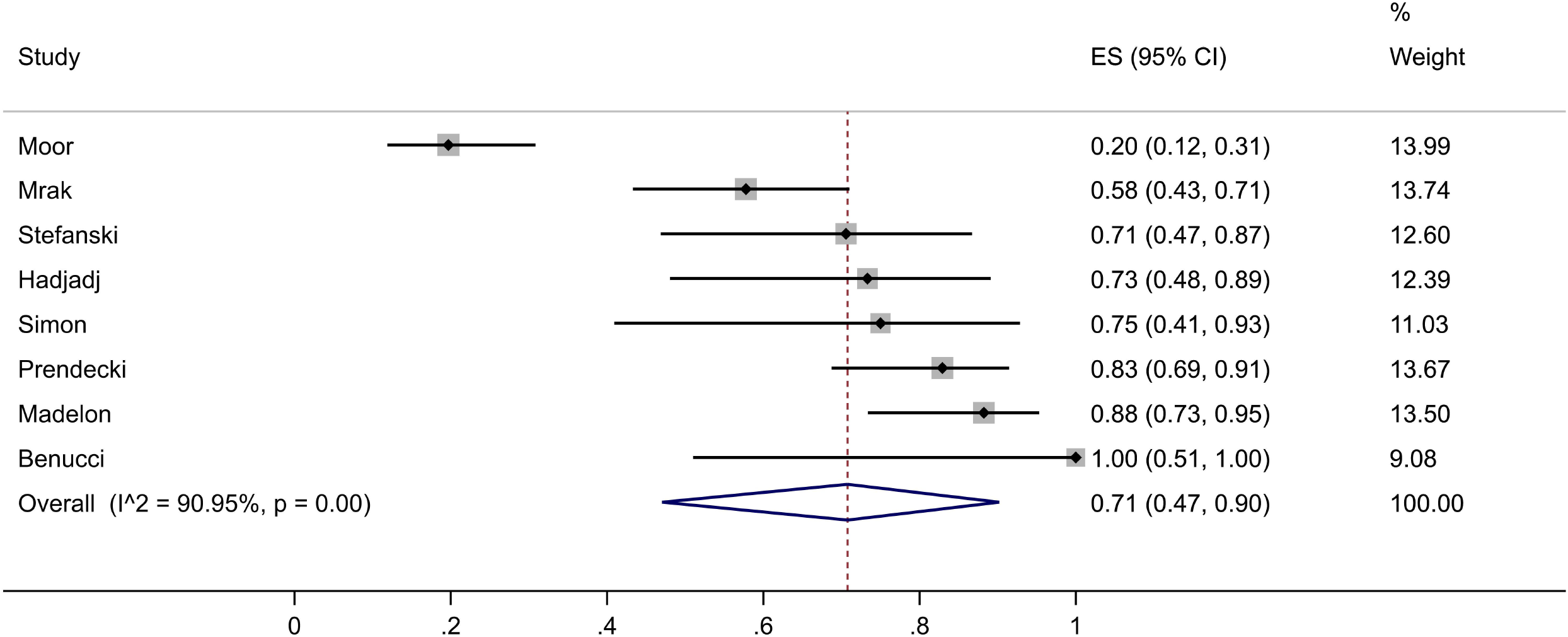
Cell-mediated immune responses across all included studies. ES, effect size. CI, confidence interval.

To address this, we next stratified response rates by assay type **(Figure 6)**. EliSpot showed a mean response rate of 72%, compared with only 22% in IGRA. However, for IGRA, results from only two studies were available, which not only differed in CMI response rates (20 vs. 100%), but also in patient numbers (66 vs. 4). Activation induced marker (AIM) analysis showed a pooled positive response rate of 83%.

**Figure 6.**
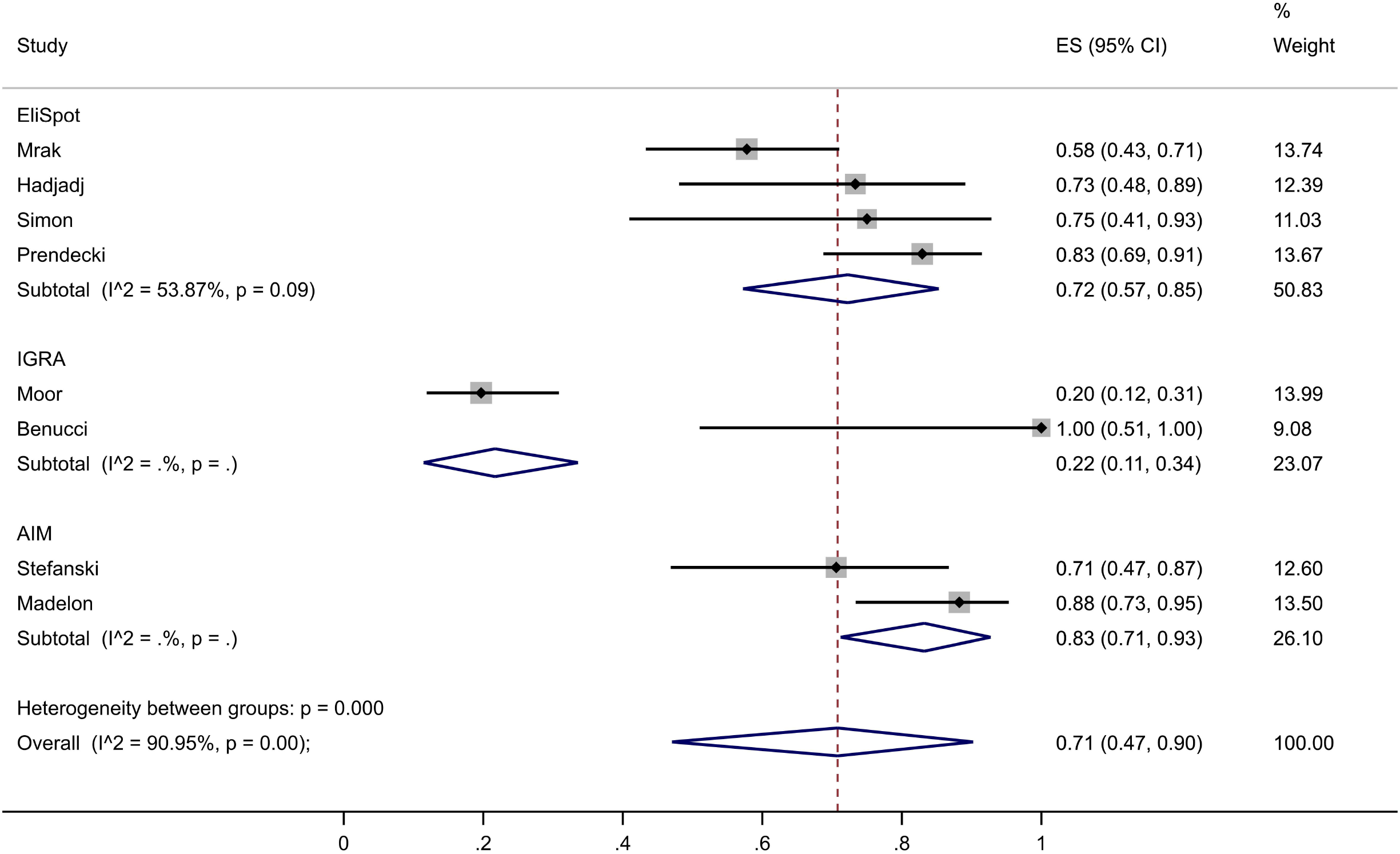
Cell-mediated immune responses according to pre-specified subgroups of assay type. IGRA, interferon-□ release assay. AIM, activation-induced marker. CI, confidence interval.

### Sensitivity analyses and reporting bias

No sensitivity analyses were conducted. There was no indication of small study effects in the Funnel plot (p-value of test for assymetry: 0.617) **(Figure 7)**.

**Figure 7.**
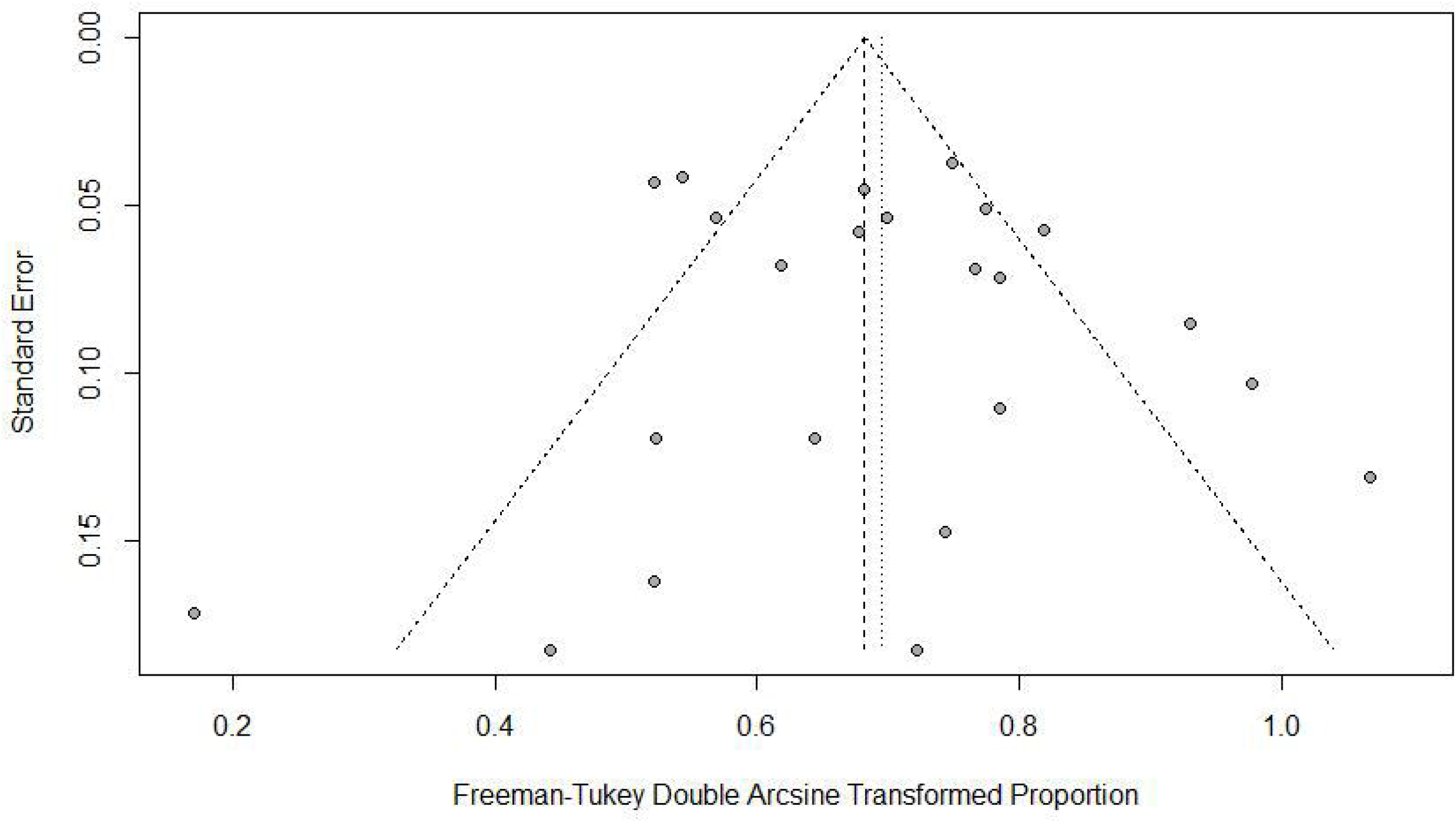
Funnel plot of all included studies.

## DISCUSSION

The present work provides an overview of seroconversion rates and cell-mediated immune responses after SARS-CoV2 vaccines from the first two dozens of studies of patients with a history of anti-CD20 therapies. Currently, no systematic reviews are available for this topic.

Our analysis suggests that a remarkable heterogeneity in immunogenicity of SARS-CoV2 vaccines exists, which partly results from differences in treatment indications, i.e. underlying underlying disease settings.

A key result is our finding that time since last anti-CD20 therapy administration severely impacts seroconversion rates. This is congruent with the results of individual studies of SARS-CoV2 vaccines ^9,10,26,38^ and has been found for influenza seroconversion in anti-CD20 therapies ^4^. This finding may be of particular interest for ideal scheduling of vaccination. However, as we have previously reported, some substantial SARS-CoV2 vaccine-induced seroconversion rates can occur in patients with high CD4-positive T cell counts even when treated with anti-CD20 therapy within the last 6 months ^10^. Further, we found that the different assays used in studies of cell-mediated immunity led to heterogeneous results. This highlights a general difference between quantitative IGRA and the more semi-quantitative EliSpot analysis, i.e. whenever an EliSpot yields a result comparable between patients and healthy controls, only the fraction of activated cells but not their quantitative activation is captured. Similar to our observation, EliSpot was reported to be more sensitive for diagnosing tuberculosis than quantitative IGRA ^40^ with an unexplained discordance between the two assay types. Moreover, we report that the type of patient collective heavily influenced the observed immune responses against SARS-CoV2. Finally, within a timeframe of 2 months after the last vaccine dose was administered, the present analysis found that seroconversion rates were not significantly changed.

The evidence base included in this review contains some limitations. First, some participants may have been included who had asymptomatic COVID19 which was not detected by serological testing e.g. by anti-nucleocapsid immunoassays. Second, the seroconversion itself is a somewhat arbitrary outcome which is heterogeneous due to manufacturer cut-offs and no clear threshold for protective antibody levels exists to date. Further, the seroconversion may not ultimately translate to protection from severe COVID19 or symptomatic COVID19 directly in patients with a history of anti-CD20 therapies. Therefore, the scarce available data on cell-mediated immunity was included in the present analysis, which represents a second albeit assay-dependent measure of immunity against SARS-CoV2.

Not enough data to allow disease subtype analysis of cellular immune responses. Published information was insufficient to allow analysis of different anti-CD20 therapies. Finally, for a closer discrimination of different autoimmune diseases, the current data were insufficient.

The present review process was further limited by an arbitrary timing of the literature search (August 21, 2021) of a rapidly emerging knowledge database which renders the current evidence preliminary rather than definitive. Further, no external experts in the field were consulted, and no unpublished studies or clinical study registry data were queried. Some potential sources of heterogeneity in seroconversion rates in the study population were not captured in subgroup analyses, such as the immunosuppressive co-medication ^10^, B cell count ^10,41^, and potentially circulating CD4-positive T cell counts in patients on anti-CD20 therapy ^10^ similar as in HIV studies ^42^. Finally, we did not perform an analysis of seroconversion rates according to the different vaccines administered, as population-level data did not sufficiently discriminate between vaccine types. Such analyses and are warranted in further studies of the evidence base.

### Summary and clinical implications

The present analysis establishes several implications for clinical practice and future research. Patients with experience of anti-CD20 therapies can and should be vaccinated against SARS-CoV2, because this successfully induces humoral and cell-mediated immune responses to SARS-CoV2 in 41% and 71% of patients, respectively. However, due to heterogeneous rates of humoral and cellular immunogenicity, patients with a treatment history of anti-CD20 therapies should be individually assessed for a personalized vaccination strategy against SARS-CoV2. While the immunogenicity of booster vaccines against SARS-CoV2 remains to be determined, we recommend a close assessment of vaccine-induced seroconversion in patients on anti-CD20 therapy for consideration of additional doses of SARS-CoV2 vaccine, especially in those within 6 months since the last dose of anti-CD20 therapy and in transplant recipients treated with multiple immunosuppressive co-medications. Finally, a threshold of at least 4 weeks after the last dose of SARS-CoV2 vaccine tends to increase seroconversion rates, which may affect future studies assessing SARS-CoV2 vaccine seroconversion in this population.

## Data Availability

All relevant data are either available in manuscript or in the appendix. Raw data and code can be requested from the corresponding author.

## Acknowledgements

The present study was funded by Bern University Hospital. The funder had no role in the design, execution of the study, analysis of the data nor decision to submit the manuscript for publication.

## Author contributions

MBM and DS conceived the study. SS, MA and MBM screened and selected studies. AL performed statistical analysis. SS, AB, MA and MBM assessed risk of bias. SS, MA, BM, MPH, DS, AB, BM, CH, DS and MBM interpreted the data. SS, MA and MBM wrote the manuscript. All authors critically reviewed and approved the final manuscript.

