## Supplementary Appendix for "Humoral and cellular immune responses upon SARS-CoV-2 vaccines in patients with anti-CD20 therapies: A systematic review and meta-analysis of 1342 patients"

**Table of contents**

Supplementary Figure	p. 2

Supplementary Table 1: Screened studies	pp. 3-7

Supplementary Table 2: Supplementary information of included studies	pp. 8-9

Supplementary Table 3: Risk of bias analysis	p. 10

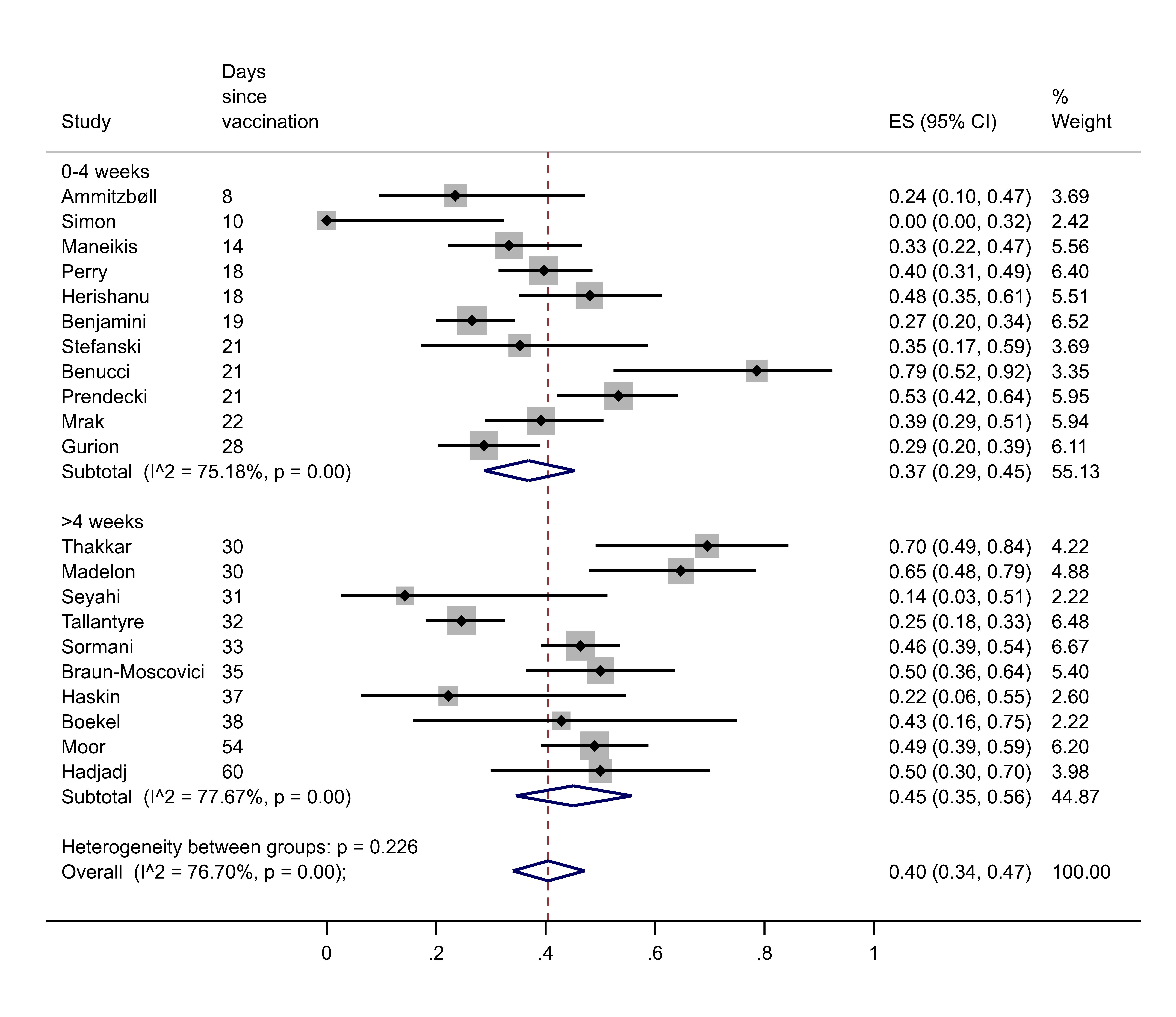

Supplementary Figure:

Humoral immune responses are indicated according to pre-specified subgroups of time > or < 4 weeks since completion of vaccination. ES, effect size; CI, confidence interval.

| Supplementary Table 1: List of screened studies | | | | |
| --- | --- | --- | --- | --- |
| **First author** | **Inclu-ded?** | **Exclu-sion stage** | **Exclusion reason** | **DOI** |
| Ammitzbøll | Y | NA | - | 10.1002/acr2.11299 |
| Benjamini | Y | NA | - | 10.3324/haematol.2021.279196 |
| Benucci | Y | NA | - | 10.1007/s12026-021-09212-5 |
| Bigaut | Y | NA | - | 10.1016/j.neurol.2021.05.001 |
| Boekel | Y | NA | - | 10.1016/S2665-9913(21)00222-8 |
| Braun-Moscovici | Y | NA | - | 10.1136/annrheumdis-2021-220503 |
| Furer | Y | NA | - | 10.1136/annrheumdis-2021-220647 |
| Gurion | Y | NA | - | I: 10.3324/haematol.2021.279216 |
| Hadjadj | Y | NA | - | 10.1101/2021.08.08.21261766 |
| Haskin | Y | NA | - | 10.1097/TP.0000000000003922 |
| Herishanu | Y | NA | - | 10.1182/blood.2021011568 |
| Madelon | Y | NA | - | 10.1101/2021.07.21.21260928 |
| Maneikis | Y | NA | - | 10.2139/ssrn.3830359 |
| Moor | Y | NA | - | 10.1101/2021.07.04.21259848 |
| Mrak | Y | NA | - | 10.1136/annrheumdis-2021-220781 |
| Perry | Y | NA | - | 10.1182/bloodadvances.2021005094 |
| Prendecki | Y | NA | - | 10.1136/annrheumdis-2021-220626 |
| Seyahi | Y | NA | - | 10.1007/s00296-021-04910-7 |
| Simon | Y | NA | - | 10.1002/art.41914 |
| Sormani | Y | NA | - | 10.2139/ssrn.3886420 |
| Stefanski | Y | NA | - | 10.1101/2021.07.19.21260803 |
| Tallantyre | Y | NA | - | 10.1101/2021.07.31.21261326 |
| Thakkar | Y | NA | - | 10.1016/j.ccell.2021.06.002 |
| Etemadifar | N | abstract | incomplete vaccina-tion | 10.1080/21645515.2021.1928463 |
| Ceccarelli | N | abstract | n ≤ 3 | 10.1177/09612033211027940 |
| Ferguson | N | abstract | n ≤ 3 | 10.1016/j.ijid.2021.06.054 |
| Wolf | N | abstract | n ≤ 3 | 10.2139/ssrn.3825910 |
| Abdelghani | N | abstract | off topic | NA |
| Agarwal | N | abstract | off topic | 10.2139/ssrn.3762626 |
| Bochnia-Bueno | N | abstract | off topic | 10.1101/2021.08.04.21261618 |
| Bochnia-Bueno | N | abstract | off topic | 10.2139/ssrn.3831124 |
| Cohen | N | abstract | off topic | 10.1007/s00259-021-05389-x |
| Costamagna | N | abstract | off topic | 10.1007/s00415-020-10149-2 |
| Daoussis | N | abstract | off topic | 10.1007/s00296-021-04969-2 |
| de Bruijn | N | abstract | off topic | 10.1111/jth.15418 |
| Drouin | N | abstract | off topic | 10.3390/v13071202 |
| Hensen | N | abstract | off topic | 10.2139/ssrn.3781631 |
| Iannetta | N | abstract | off topic | 10.1016/j.msard.2021.103157 |
| Kant | N | abstract | off topic | 10.1016/j.ekir.2021.03.876 |
| Klineova | N | abstract | off topic | 10.1016/j.msard.2021.103153 |
| Kovvuru | N | abstract | off topic | 10.2139/ssrn.3633220 |
| Machado | N | abstract | off topic | NA |
| Margoni | N | abstract | off topic | 10.1007/s00415-021-10744-x |
| Mateen | N | abstract | off topic | 10.1007/s00415-020-10045-9 |
| Meng Poh | N | abstract | off topic | 10.1038/s41467-020-16638-2 |
| Pedotti | N | abstract | off topic | 10.1016/j.msard.2021.103203 |
| Penner | N | abstract | off topic | 10.2139/ssrn.3798557 |
| Riches | N | abstract | off topic | 10.5501/wjv.v10.i3.97 |
| Rizk | N | abstract | off topic | 10.1001/jamacardio.2021.3444 |
| Sellebjerg | N | abstract | off topic | 10.1097/WCO.0000000000000938 |
| Shakoor | N | abstract | off topic | 10.1053/j.ajkd.2021.06.016 |
| Swaims-Kohlmeier | N | abstract | off topic | 10.2139/ssrn.3806678 |
| Takuva | N | abstract | off topic | 10.2139/ssrn.3845988 |
| Tzarnas | N | abstract | off topic | NA |
| Valtanen | N | abstract | off topic | 10.2139/ssrn.3859298 |
| Waldman | N | abstract | off topic | 10.1016/j.jaad.2020.10.075 |
| Arnold | N | abstract | review | 10.1093/rheumatology/keab223 |
| Baker | N | abstract | review | 10.1111/cei.13495 |
| Barrière | N | abstract | review | 10.1016/j.ejca.2021.06.008 |
| Bhise | N | abstract | review | 10.1007/s13311-021-01008-7 |
| Bruchfeld | N | abstract | review | 10.1093/ndt/gfab174 |
| Cabreira | N | abstract | review | 10.3390/vaccines9070773 |
| Ciotti | N | abstract | review | 10.1016/j.msard.2020.102439 |
| Coyle | N | abstract | review | 10.1007/s12325-021-01761-3 |
| Houot | N | abstract | review | 10.1016/j.ejca.2020.06.017 |
| Inshasi | N | abstract | review | 10.1007/s40120-021-00260-5 |
| Kelly | N | abstract | review | 10.1016/j.jneuroim.2021.577599 |
| Mason | N | abstract | review | 10.1177/09612033211024355 |
| Otero-Romero | N | abstract | review | 10.1097/WCO.0000000000000929 |
| Park | N | abstract | review | 10.3346/jkms.2021.36.e95 |
| Santosa | N | abstract | review | 10.1111/1756-185X.14107 |
| Soy | N | abstract | review | 10.1007/s10067-021-05700-z |
| Sparks | N | abstract | review | 10.1097/BOR.0000000000000812 |
| Vijenthira | N | abstract | review | 10.1182/bloodadvances.2021004629 |
| Chilimuri | N | abstract | SARS-CoV2 infection status | 10.1093/rap/rkab038 |
| Duléry | N | abstract | SARS-CoV2 infection status | 10.1002/ajh.26209 |
| Flores-Gonzalez | N | abstract | SARS-CoV2 infection status | 10.1016/j.msard.2021.102777 |
| Mantus | N | abstract | SARS-CoV2 infection status | 10.4049/jimmunol.2001420 |
| Stastna | N | abstract | SARS-CoV2 infection status | 10.1016/j.msard.2021.103104 |
| Ammitzbøll | N | duplicate check | duplicate | 10.1002/acr2.11299 |
| Ammitzbøll | N | duplicate check | duplicate | 10.1002/acr2.11299 |
| Arnold | N | duplicate check | duplicate | 10.1093/rheumatology/keab223 |
| Baker | N | duplicate check | duplicate | 10.1111/cei.13495 |
| Baker | N | duplicate check | duplicate | 10.1111/cei.13495 |
| Benjamini | N | duplicate check | duplicate | 10.3324/haematol.2021.279196 |
| Bigaut | N | duplicate check | duplicate | 10.1016/j.neurol.2021.05.001 |
| Bochnia-Bueno | N | duplicate check | duplicate | 10.2139/ssrn.3831124 |
| Boekel | N | duplicate check | duplicate | 10.2139/ssrn.3869656 |
| Boekel | N | duplicate check | duplicate | 10.2139/ssrn.3869656 |
| Brosh-Nissimov | N | duplicate check | duplicate | 10.1016/j.cmi.2021.06.036 |
| Bruchfeld | N | duplicate check | duplicate | 10.1093/ndt/gfab174 |
| Cabreira | N | duplicate check | duplicate | 10.3390/vaccines9070773 |
| Chilimuri | N | duplicate check | duplicate | 10.1093/rap/rkab038 |
| Cohen | N | duplicate check | duplicate | 10.1007/s00259-021-05389-x |
| Cohen | N | duplicate check | duplicate | 10.1007/s00259-021-05389-x |
| Coyle | N | duplicate check | duplicate | 10.1007/s12325-021-01761-3 |
| Coyle | N | duplicate check | duplicate | 10.1007/s12325-021-01761-3 |
| de Brujin | N | duplicate check | duplicate | 10.1111/jth.15418 |
| Deepak | N | duplicate check | duplicate | 10.1101/2021.04.05.21254656 |
| Drouin | N | duplicate check | duplicate | 10.3390/v13071202 |
| Etemadifar | N | duplicate check | duplicate | 10.1080/21645515.2021.1928463 |
| Furer | N | duplicate check | duplicate | 10.1136/annrheumdis-2021-220647 |
| Furer | N | duplicate check | duplicate | 10.1136/annrheumdis-2021-220647 |
| Gurion | N | duplicate check | duplicate | I: 10.3324/haematol.2021.279216 |
| Herishanu | N | duplicate check | duplicate | 10.1182/blood.2021011568 |
| Herzog Tsarfati | N | duplicate check | duplicate | [10.1002/ajh.26284](https://doi.org/10.1002/ajh.26284) |
| Houot | N | duplicate check | duplicate | 10.1016/j.ejca.2020.06.017 |
| Inshasi | N | duplicate check | duplicate | 10.1007/s40120-021-00260-5 |
| Inshasi | N | duplicate check | duplicate | 10.1007/s40120-021-00260-5 |
| Kant | N | duplicate check | duplicate | 10.1016/j.ekir.2021.03.876 |
| Klineova | N | duplicate check | duplicate | 10.1016/j.msard.2021.103153 |
| Kovvuru | N | duplicate check | duplicate | 10.2139/ssrn.3633220 |
| Madelon | N | duplicate check | duplicate | 10.1101/2021.07.21.21260928 |
| Maneikis | N | duplicate check | duplicate | 10.2139/ssrn.3830359 |
| Maneikis | N | duplicate check | duplicate | 10.2139/ssrn.3830359 |
| Mason | N | duplicate check | duplicate | 10.1177/09612033211024355 |
| Mateen | N | duplicate check | duplicate | 10.1007/s00415-020-10045-9 |
| Park | N | duplicate check | duplicate | 10.3346/jkms.2021.36.e95 |
| Peeters | N | duplicate check | duplicate | 10.2139/ssrn.3868066 |
| Peeters | N | duplicate check | duplicate | 10.2139/ssrn.3868066 |
| Peeters | N | duplicate check | duplicate | 10.2139/ssrn.3868066 |
| Prendecki | N | duplicate check | duplicate | 10.1136/annrheumdis-2021-220626 |
| Rizk | N | duplicate check | duplicate | 10.1001/jamacardio.2021.3444 |
| Santosa | N | duplicate check | duplicate | 10.1016/j.jneuroim.2021.577599 |
| Santosa | N | duplicate check | duplicate | 10.1111/1756-185X.14107 |
| Santosa | N | duplicate check | duplicate | 10.1101/2021.03.01.21252653 |
| Sellebjerg | N | duplicate check | duplicate | 10.1097/WCO.0000000000000938 |
| Sellebjerg | N | duplicate check | duplicate | 10.1097/WCO.0000000000000938 |
| Seyahi | N | duplicate check | duplicate | 10.1007/s00296-021-04910-7 |
| Shakoor | N | duplicate check | duplicate | 10.1053/j.ajkd.2021.06.016 |
| Sormani | N | duplicate check | duplicate | 10.2139/ssrn.3886420 |
| Sparks | N | duplicate check | duplicate | 10.1097/BOR.0000000000000812 |
| Stefanski | N | duplicate check | duplicate | 10.1101/2021.07.19.21260803 |
| Stefanski | N | duplicate check | duplicate | 10.1101/2021.07.19.21260803 |
| von Lilienfeld-Toal | N | duplicate check | duplicate | 10.1007/s00761-021-00972-1 |
| Waldman | N | duplicate check | duplicate | 10.1016/j.jaad.2020.10.075 |
| Peeters | N | full text | information missing | 10.2139/ssrn.3868066 |
| Herzog Tsarfati | N | full text | n ≤ 3 | [10.1002/ajh.26284](https://doi.org/10.1002/ajh.26284) |
| Haroun | N | full text | off topic | NA |
| Parry | N | full text | responder numbers not specified | 10.2139/ssrn.3845994 |
| Deepak | N | full text | responder numbers not specified | 10.1101/2021.04.05.21254656 |
| Maneikis | N | full text | responder numbers not specified | 10.1016/S2352-3026(21)00169-1 |
| Conte | N | full text | SARS-CoV2 infection status | NA |
| Cook | N | full text | SARS-CoV2 infection status | 10.1101/2021.08.04.21261618 |
| Sottini | N | full text | SARS-CoV2 infection status | NA |
| Wallach | N | full text | SARS-CoV2 infection status | 10.1016/j.msard.2021.102793 |
| Brosh-Nissimov | N | full text | SARS-CoV2 infection status | 10.1016/j.cmi.2021.06.036 |

Y, Yes. N, No. NA, not available

| Supplementary table 2: Supplementary information of included studies | | | | | |
| --- | --- | --- | --- | --- | --- |
| **First author** | **Time from vaccine to serology** | **Time (days) from vaccine to assessment** | **AB Cut-off** | **CMI cut-off** | Anti-CD20 monotherapy |
| Boekel | >4w | 38 | cut-off 4 AU/ml | NA | NA in population-level data |
| Perry | 0-4w | 17.5 | >0.80 U/mL considered to be positive. | NA | NA in population-level data |
| Haskin | >4w | 37 | >50AU/ml | NA | None |
| Prendecki | 0-4w | 21 | 7.1 BAU/ml | cut- off for positivity of 40 SFU/106 PBMC for S protein | NA in population-level data |
| Herishanu | 0-4w | 17.5 | >0.80 U/mL considered to be positive. | NA | NA in population-level data |
| Gurion | 0-4w | 28 | 50AU/ml | NA | NA in population-level data |
| Benjamini | 0-4w | 19 | * 1) >15 units/mL; 2) >50 u/mL; 3) >1.1 | NA | NA in population-level data |
| Ammitzbøll | 0-4w | 8 | >1.0 index (s/c) | NA | Humoral responders 3/10 (30%) |
| Bigaut | NA | NA | threshold between 0.72 and 1.54 U/ ml | NA | NA in population-level data |
| Thakkar | >4w | 30 | >50 arbitrary units per milliliter [AU/mL] | NA | NA in population-level data |
| Furer | NA | NA | above 15 binding antibody units (BAU) | NA | Humoral responders 11/28 (39%) |
| Seyahi | >4w | 30.7 | ≥ 0.8 U/mL | NA | NA in population-level data |
| Benucci | 0-4w | 21 | >10U/ml | NA | None |
| Mrak | 0-4w | 21.9 | Quantitation range 0.4-2500.0 U/mL. |  | NA in population-level data |
| Simon | 0-4w | 10 | > 1.1 index |  | NA in population-level data |
| Braun-Moscovici | >4w | 35 | positive above 50 AU/mL | NA | NA in population-level data |
| Hadjadj | >4w | 60 | antibody binding unit (BU) above 1.1 for IgG and 0.2 for IgA |  | NA in population-level data |
| Madelon | >4w | 30 | anti-RBD; 0.8 U/ml |  | NA in population-level data |
| Stefanski | 0-4w | 21 | > 1.1 index |  | NA in population-level data |
| Tallantyre | >4w | 32.2 | ≥0.7 INDEX, RBP | NA | NA in population-level data |
| Moor | >4w | 53.7 | > 1.1 index | >15mU/ml | Humoral responders 27/39 (69%), cellular 10/27 (37%) |
| Maneikis | 0-4w | 14 | 50AU/ml | NA | NA in population-level data |
| Sormani | >4w | 33 | 0.80 U/mL for RBD | NA | NA in population-level data |
| *assay: 1 of 3 kits: 1) Liaison S1/S2 IgG (Diasorin); 2) Architect AdviseDx (Abbott); 3) RBD-ELISA | | | | | |

| Supplementary table 3: Risk of bias analysis | | | | |
| --- | --- | --- | --- | --- |
| Study | Selection | Comparability | Outcome | Overall Risk of Bias |
| Boekel | ******** | ***** | ****** | **Low**-Medium |
| Perry | ******** | ***** | ****** | **Low**-Medium |
| Haskin | ******* | ***** | ******* | **Low**-Medium |
| Prendecki | ******** | ***** | ****** | **Low**-Medium |
| Herishanu | ******* | ***** | ****** | **Low**-Medium |
| Gurion | ****** | ***** | ****** | Low-**Medium** |
| Benjamini | ******* | ***** | ****** | **Low**-Medium |
| Ammitzbøll | ******* | ****** | ****** | Low |
| Bigaut | ****** | ***** | ****** | Low-**Medium** |
| Thakkar | ****** | ***** | ****** | Low-**Medium** |
| Furer | ****** | ****** | ***** | Low-**Medium** |
| Seyahi | ****** | ***** | ****** | Low-**Medium** |
| Benucci | ***** | ***** | ***** | **Medium**-High |
| Mrak | ***** | ****** | ***** | **Medium**-High |
| Simon | ******* |  | ***** | Medium |
| Braun-Moscovici | ******* | ***** | ****** | **Low**-Medium |
| Hadjadj | ******* |  | ****** | Low-**Medium** |
| Madelon | ****** |  | ****** | Medium |
| Stefanski | ******** | ***** | ***** | Low-**Medium** |
| Tallantyre | ******** | ***** | ****** | **Low**-Medium |
| Moor | ******* | ****** | ****** | Low |
| Maneikis | ******* | ***** | ***** | Low-**Medium** |
| Sormani | ******* | ***** | ****** | **Low**-Medium |
| Boekel | ******** | ***** | ****** | **Low**-Medium |
| Perry | ******** | ***** | ****** | **Low**-Medium |
| Haskin | ******* | ***** | ******* | **Low**-Medium |
| Prendecki | ******** | ***** | ****** | **Low**-Medium |

**Scoring used for Newcastle-Ottawa assessment tool**

| **Domain** | **Good Quality/Low Risk** | **Fair Quality/Medium Risk** | **Low Quality/High Risk** |
| --- | --- | --- | --- |
| Selection | 3-4 stars | 2 stars | 0-1 stars |
| Comparability | 2 stars | 1 star | 0 stars |
| Outcome | 2-3 stars | 1 star | 0 stars |

For mixed ratings, i.e. “low-medium”, the overall rating is closer to the **bold-marked** rating
